# Long-Read Sequencing Increases Diagnostic Yield for Pediatric Sensorineural Hearing Loss

**DOI:** 10.1101/2024.09.30.24314377

**Authors:** Shelby E. Redfield, Wanqing Shao, Tieqi Sun, Adrian Pastolero, William J. Rowell, Courtney E. French, Cillian Nolan, J. Matthew Holt, Christopher T. Saunders, Cairbre Fanslow, Eirini Maria Lampraki, Christine Lambert, Margaret Kenna, Michael Eberle, Shira Rockowitz, Aiden Eliot Shearer

## Abstract

The diagnostic yield of genetic testing for pediatric sensorineural hearing loss (SNHL) has remained at around 40% for over a decade despite newly discovered causative genes and the expanded use of exome sequencing (ES). This stagnation may be due to (1) a focus on coding regions of the genome and (2) an inability to resolve variants in complex genomic regions due to reliance on short-read sequencing technologies. Short-read genome sequencing (srGS) and long-read genome sequencing (lrGS) both provide exonic single nucleotide variant (SNV) and small indel detection at the same sensitivity as ES, but also evaluate intronic regions. lrGS provides improved resolution for structural variants (SV) and repetitive genomic regions. We sought to investigate the potential utility of lrGS in the diagnostic evaluation of a small cohort of patients with SNHL of unknown etiology after ES and srGS. 19 pediatric patients with SNHL underwent lrGS via PacBio SMRT sequencing. Sequencing data were processed using the PacBio WGS variant pipeline. The diagnostic yield for this lrGS cohort was 4/19 (21%). Relevant variants detected only with lrGS included a hemizygous deletion *in trans* with a missense variant in an area of high genomic homology (*OTOA*) and two single nucleotide loss-of-function variants *in trans* to a known copy-number-loss for a gene with a highly homologous pseudogene (*STRC)*. A complex inversion was identified in the *MITF* gene which was also identified on post-hoc analysis by srGS. LrGS provides improved resolution for complex genomic structural variation which may increase diagnostic yield for genetic pediatric SNHL, and, potentially, rare disease more broadly.

## Introduction

Congenital sensorineural hearing loss (SNHL) affects as many as 1 in 500 infants in the United States (Morton and Nance 2006) and a majority of these cases are attributable to a Mendelian genetic etiology (Smith et al. 2005). The implementation of comprehensive genetic testing for pediatric SNHL, typically via targeted SNHL gene panels or exome sequencing (ES), results in an overall diagnostic yield of approximately 43% (Downie et al. 2020; Perry et al. 2023; Rouse et al. 2022; Shearer 2024; Sloan-Heggen et al. 2016). There is variation in yield based on clinical phenotype and in some populations, for instance congenital severe to profound hearing loss, the diagnostic yield can approach 60% (Downie et al. 2020; Sloan-Heggen et al. 2016). This diagnostic yield is relatively high compared to other neurodevelopmental disorders for which genetic testing is routinely obtained, such as autism (17%) and epilepsy (24%) (Stefanski et al. 2021). However, this means that up to 60% of children with a high *a priori* suspicion of a hereditary SNHL still go undiagnosed (Kim et al. 2022; Retterer et al. 2016).

The most commonly applied testing methodology for the genetic diagnosis of SNHL is targeted gene panels, which rely on short read (150-300bp) sequencing of a targeted list of SNHL genes. Less frequently, pediatric patients with a primary indication of SNHL will undergo ES, also using short-read sequencing technologies. Both gene panels and ES are limited by (1) a focus on exonic or coding regions and nearby splice sites, and (2) low resolution for complex structural variation that contributes to genetic disease. Short-read genome sequencing (srGS), which is currently not commonly applied in the clinical evaluation of individuals with SNHL, does provide effective coverage of intronic and noncoding regions (Byrska-Bishop et al. 2022; Pagnamenta et al. 2023).

However, while srGS evaluates noncoding regions of the genome, it is often unable to resolve areas of low complexity, high duplication rate, and structural variation (Chaisson et al. 2019; Genomes Project et al. 2015). In fact, much of the human genome maps to segmental duplications, or stretches of DNA over 1kb long with over 90% sequence identity, that are highly mutable but very challenging to resolve using current methodologies (Bailey et al. 2001; Vollger et al. 2023). Two genes that commonly cause nonsyndromic SNHL, *OTOA* and *STRC*, map to a segmental duplication and highly homologous pseudogene, respectively, and detecting pathogenic variants within these genes is challenging (Abbasi et al. 2022; Shearer et al. 2014). This is particularly important given that pathogenic variation in *STRC* is the second most common cause of genetic SNHL (Shearer et al 2014).

Long-read genome sequencing (lrGS), by contrast, generates sequencing reads up to several megabases long (Mahmoud et al. 2024; Mantere et al. 2019) and therefore has high resolution for complex structural variation and repetitive genomic regions. Several studies over the last few years have shown that long-read techniques have the capacity to identify Mendelian etiology after standard sequencing techniques have failed, including a handful of patients with SNHL (Conlin et al. 2022; Miller et al. 2021; Ohori et al. 2021; Olivucci et al. 2024; Vache et al. 2020; Xie et al. 2020).

In this study, our goal was to evaluate the potential utility of lrGS for genetic diagnosis for pediatric hearing loss in patients with nondiagnostic ES and srGS. We find that lrGS provided significantly improved resolution for complex structural variation and, in this cohort, substantially improved diagnostic yield over ES and srGS.

## Methods

### Recruitment and Participants

This research was approved by the Institutional Review Board at Boston Children’s Hospital (P00035179 and P00031494) and performed in partnership with the Children’s Rare Disease Cohort initiative. Pediatric patients with SNHL were recruited in the Boston Children’s Hospital Otolaryngology and Communication Enhancement clinical setting and provided written informed consent to participate. Criteria for eligibility included having a clinical diagnosis of SNHL that did not have a known genetic etiology; there were no exclusions based on other clinical criteria, including age of onset, laterality or severity of hearing loss, presence of syndromic features, or family history. All patients had diagnostic work up for sensorineural hearing loss including audiometric testing and clinical evaluation; many had additional diagnostic workup including congenital cytomegalovirus (cCMV) testing and temporal bone imaging (CT and/or MRI).

### Genetic Testing Methodologies

Prior to lrGS through this study, genetic testing methodologies varied based on individual patient circumstances. Some probands had prior single gene sequencing or targeted hearing loss gene panel sequencing on a clinical basis. All probands had research exome sequencing, and most probands had srGS performed on a research basis prior to research lrGS. Prior genetic testing, regardless of methodology, was deemed nondiagnostic by the testing laboratory and clinical team.

### DNA Extraction, Sequencing, and Analysis

DNA was extracted from whole blood using standard methods. 150bp paired end ES and srGS was performed at GeneDx using Illumina sequencers. ES and srGS data were processed using Illumina Dragen v3.9 as previously described (Rockowitz et al. 2020). For PacBio sequencing, 12-15 kb insert SMRTbell libraries were prepared using the SMRTbell prep kit 3.0. Long-read whole genome sequencing was performed using Single Molecule, Real-Time (SMRT) sequencing at Pacific Biosciences (“PacBio,”) (Menlo Park, CA) using the PacBio Sequel II system to a depth of 24-32x coverage. Sequence data was processed by PacBio variant pipeline. Unaligned HiFi reads in BAM format were received from PacBio. Read alignment and variant calling were performed using PacBio HiFi-human-WGS-WDL workflow v1.0.3. In short, HiFi reads were aligned to the GRCh38 (GCA_000001405.15) genome using pbmm2 v1.10.0. Single nucleotide variants, small insertions and deletions were identified with DeepVariant v1.5.0 (Poplin et al. 2018). Structural variants were called with pbsv v2.9.0. Copy number variations were called with HiFiCNV v0.1.7. Short tandem repeat expansion was identified using TRGT v0.5.0. Small variants and structural variants were jointly phased with HiPhase v0.10.2 (Holt et al. 2024), and phase haplotype tags (“HP”) were added to the aligned BAM file. Variant calling results were further processed using in house bioinformatic pipelines and analyzed via GeneDx Discovery Platform. For each candidate variant, sequencing reads were visually reviewed using IGV v2.16.0.

### Clinical Confirmation and Return of Results

Variant pathogenicity was determined according to the American College of Medical Genetics and the Associations for Molecular Pathology guidelines as modified for SNHL-specific considerations (Oza et al. 2018; Richards et al. 2015). Candidate variants that were identified via research analysis of lrGS were clinically confirmed using an additional clinical sample and orthogonal genetic testing method within a commercial genetic testing laboratory (Prevention Genetics, Marshfield, WI) environment with Clinical Laboratory Improvement Amendments (CLIA) approval. Clinically confirmed variant results were returned to families and genetic counseling was provided by a certified and licensed genetic counselor and otolaryngologist.

## Results

### Subject Demographic and Clinical Characteristics

Nineteen probands underwent lrGS through this study (**Table 1**). The median age at the time of results was 7-years-old (with an interquartile range of 5.1-years-old to 9.7-years-old). Fifteen (79%) probands had a bilateral symmetric SNHL, three (15.8%) had bilateral asymmetric hearing loss, and one (5.2%) had unilateral SNHL with bilateral enlarged vestibular aqueducts. Twelve probands (63%) had congenital onset SNHL, 2 (10.5%) had prelingual SNHL, 3 (15.8%) had postlingual SNHL, and onset of SNHL was unknown for 2 probands (10.5%). Severity of SNHL was highly variable (**Supplemental Table 1**). Two (10.5%) probands had an autosomal recessive family history of SNHL, two (10.5%) had an autosomal dominant family history of SNHL, and 15 (78.9%) had no reported family history. Five (33.3%) probands had notable extra-auditory clinical presentations including hemolytic anemia; retinal dystrophy; global developmental delay; hypotonia, low vision, and nystagmus, and hematuria in one proband each.

**Table 1:** Cohort demographic and clinical characteristics. lrGS: long-read genome sequencing.

| Characteristic | IrGS Cohort n (%) |
| --- | --- |
| **Total** | 19 (100%) |
| **Sex** |  |
| Female | 13 (68.4%) |
| Male | 6 (31.6%) |
| **Race** |  |
| White | 16 (82.4%) |
| Not Reported | 1 (5.2%) |
| Asian | 1 (5.2%) |
| Hispanic/Latino | 1 (5.2%) |
| **Positive Family History (of childhood-onset HL)** |  |
| No | 15 (78.9%) |
| Yes | 4 (21.1%) |
| **HL Onset** |  |
| Congenital | 12 (63.1%) |
| Prelingual | 2 (10.5%) |
| Postlingual | 3 (15.8%) |
| Unknown | 2 (10.5%) |
| **HL Laterality** |  |
| Bilateral, symmetric | 15 (78.9%) |
| Unilateral | 1 (5.2%) |
| Bilateral, asymmetric | 3 (15.8%) |
| **HL Severity (in worse ear)** |  |
| Mild-Moderate | 9 (47.4%) |
| Severe-Profound | 10 (52.6%) |
| **Syndromic Features** |  |
| Yes | 5 (26.3%) |
| No | 14 (73.7%) |
| **Previous Genetic Testing** |  |
| Single gene | 3 (15.8%) |
| Gene panel | 4 (21.1%) |
| Exome sequencing | 19 (100%) |
| Short read genome sequencing | 18 (94.7%) |

All probands had extensive genetic testing prior to lrGS, the primary indication for which was SNHL. Three probands (15.8%) had single gene (GJB2) testing, four probands (21%) had SNHL gene panel testing, 19 probands (100%) had exome sequencing, and 18 probands (94.7%) had short read genome sequencing. Testing was nondiagnostic for all 19 probands on initial analysis. Nine probands (47.4%) were identified to harbor a heterozygous pathogenic or likely pathogenic variant in a gene that was associated exclusively or primarily with autosomal recessive conditions, including two probands with a heterozygous loss of *STRC* (full patient details are included in **Supp. Table 1**).

### Long Read Genome Sequencing

The median read length across 19 samples was 13.5 kb (1.2 kb standard deviation). The median depth of coverage of lrGS across samples was 29.5x (3.5x standard deviation). On average, per sample, 5,386,525 single nucleotide variants were detected (range 5,305,533-5,533,276) and 22,004 structural variants were detected (range 19,838-23,049). The number of variants, both SNVs and SVs, detected through ES and srGS of these same patients was significantly lower than for lrGS (**Table 2 and Supp. Table 2**).

**Table 2.** Key sequencing statistics using exome sequencing (ES), short-read genome sequencing (srGS) and long-read genome sequencing (lrGS).

| Method | ES | srGS | PacBio lrGS |
| --- | --- | --- | --- |
| N Samples | 19 | 18 | 19 |
| Read Length (bp) | 150 | 150 | 13,539 |
| Avg Depth of Coverage | 75.5 | 50.9 | 29.5 |
| SNV Detected (T-test* significance relative to lrGS) | 117,336** (p=4.2E-36) | 5,039,980 (p=1.2E-18) | 5,386,525 |
| SV Detected (T-test* significance relative to lrGS) | 78<sup>†</sup> (p=1.5E-28) | 12,273 (p=6.6E-15) | 22,004 |

SNV = single nucleotide variant, SV = structural variant. \*Two-tailed paired T-test. \*\*T-test significance ES relative to srGS for SNVs: 5.1E-34. <sup>†</sup>T-test significance ES relative to srGS for SVs: 9.6E-17.

LrGS and analysis was completed for 19 probands. Genetic variants were identified for 4 of the 19 probands that were assessed to be causative or potentially causative resulting in a diagnostic yield of 21.1%.

### Case Vignettes

#### Case 1

Proband 1 is a with a bilateral mild-moderate SNHL who is otherwise healthy. Proband referred bilaterally on the newborn hearing screen in the neonatal period. Two diagnostic auditory brainstem response (ABR) tests in infancy showed a bilateral SNHL in the mild range of severity. Subsequent behavioral audiometric evaluations in childhood have been consistent with a stable bilateral mild sensorineural hearing loss. The proband has an older sibling with a similar history and audiological profile. There is otherwise no family history of SNHL. Proband had cytomegalovirus (CMV) testing in the newborn period that was negative. She has not had diagnostic imaging of the inner ear. Research ES for proband, sibling, and parents was nondiagnostic but did identify a heterozygous copy number loss at chromosome 15q15.3 encompassing the *STRC* and *CATSPER2* genes. No second variant was detected. SrGS did not identify any further notable variants.

lrGS identified a pathogenic nonsense mutation in trans with a hemizygous deletion of *STRC* and *CATSPER2* (**Figure 1A**). The depth-based CNV caller HiFiCNV detected a 104 kb deletion (approx. chr15:43,566,001-43,670,000). On the other allele, DeepVariant (Poplin et al. 2018) analysis identified the variant NM_153700.2:c.1228C>A, p.(Gln410*) (chr15:43,616,338 G>A). This variant is absent from gnomAD, ClinVar, and LOVD and results in a stop gain in an exon that is challenging to map with short reads due to a highly similar paralog. Interestingly, phasing analysis using Paraphase (Chen et al. 2023) enabled visualization of a single *STRC* haplotype and two pseudogene haplotypes of *pSTRC* (**Figure 1A)**. This phasing analysis of a complex region helps to confirm segregation. Both *STRC* variants were clinically confirmed and disclosed to the family. LrGS was able to help resolve this SNV when other methods could not, due to the highly homologous pseudogene *pSTRC*.

**Figure 1.**
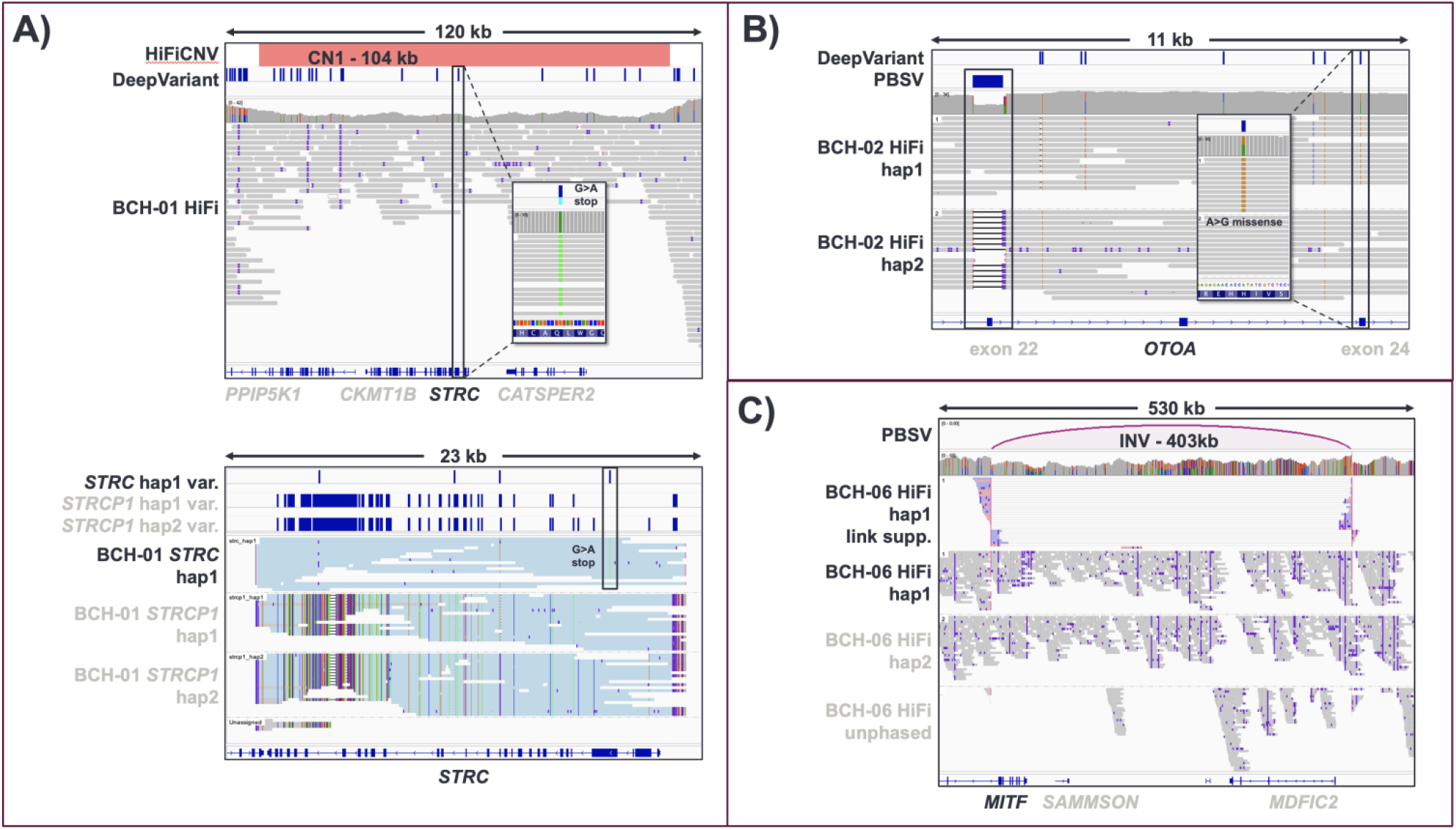
Causative variants identified via long-read genome sequencing (lrGS). A) Top panel shows identification of compound heterozygous pathogenic variant and hemizygous deletion in *STRC*. Bottom panel shows haplotype analysis indicating two *STRCP1* haplotypes and only one *STRC* haplotype, with roughly half of the expected coverage (CN1), containing a G>A stop gain at chr15:43,616,338. B) Identification of a single exon deletion (exon 22) and stop mutation in compound heterozygosity in the *OTOA* gene. C) Identification of a copy-neutral inversion in the *MITF* gene.

#### Case 2

Proband 2 is a male with a bilateral mild to moderate SNHL who is otherwise healthy. He did not pass a newborn hearing screen. An ABR performed in infancy identified a mild SNHL. A recent behavioral audiometric evaluation in childhood showed a bilateral mild to moderate SNHL. The proband had negative CMV testing in the newborn period. He has not had diagnostic imaging of the inner ear. Research trio ES identified a heterozygous CNV at 15q15.3 encompassing the *STRC* and *CATSPER2* genes. SrGS was nondiagnostic. Similar to Case 1, LrGS identified a single nucleotide variant in *STRC* (NM_153700.2:c.3217C>T, p.(Arg1073*)) *in trans* to the CNV which was classified as pathogenic. Following the identification of these *STRC* variants, another sibling to proband 2 was born and identified to have a mild bilateral SNHL shortly after birth; results of targeted STRC variant testing for this sibling are pending.

#### Case 3

Proband 3 is a female with a bilateral SNHL. She did not pass a newborn hearing screen bilaterally. Initial ABR in infancy identified a bilateral SNHL that was moderate in severity. A recent hearing test in childhood demonstrated a slightly u-shaped audiogram configuration in the moderate to moderately-severe range of severity. She had CMV testing that was negative. She has not had diagnostic imaging of the inner ear. Targeted gene panel testing identified a heterozygous likely pathogenic variant in *MYO7A* (**Supp. Table 1**.) The proband’s unaffected parent also harbored this variant. Research exome and srGS were nondiagnostic with no further variants of interest identified.

LrGS identified a copy number variant (CNV) (loss of 769bp at chr16:21,735,945) including exon 22 of *OTOA in trans* with a heterozygous missense variant (NM_144672.3:c.2654A>G, p.(His885Arg)) in the *OTOA* gene (**Figure 1B**). This result was deemed to be possibly/likely diagnostic of *OTOA-*associated nonsyndromic hearing loss. However, the missense variant in *OTOA* is currently classified as a variant of uncertain significance. *OTOA* is within a segmental duplication. Neither variant in *OTOA* had been appreciated before lrGS; upon review, the srGS analysis pipeline had called the His885Arg missense variant, but it had been filtered out of the list of candidate variants due to low call quality (poor coverage and mapping quality). The CNV was detected only by lrGS.

#### Case 4

Proband 4 is a female with a bilateral asymmetric SNHL. She passed a newborn hearing screen bilaterally. Failing a hearing screen in early childhood prompted audiological assessment. Behavioral audiometry identified a mild low-frequency SNHL, which has progressed slightly since first identified. In the right ear, hearing is in the moderate rising to normal range of severity. In the left ear, hearing has been in the slight sloping to moderate rising to normal range of severity. *GJB2* sequencing was negative. Research ES was nondiagnostic.

lrGS identified a 403.2kb inversion (chr3:69977069_70380310inv) upstream of the *MITF* gene predicted to impact alternatively spliced transcripts of *MITF*. Similar inversions have not been reported before, so this variant was classified as a variant of uncertain significance though it is suspected to be causative of the proband’s SNHL. This copy-neutral inversion was subsequently identified on srGS upon reanalysis of data using an updated tertiary analysis pipeline.

## Discussion

Despite increasing access to comprehensive genetic testing for pediatric SNHL patients, the discovery of additional SNHL genes, and decreasing costs of testing, the diagnostic yield of the genetic evaluation of SNHL remains at approximately 43% over the past decade (Shearer 2024). While this diagnostic rate is high compared to other indications for genomic testing in children (Malinowski et al. 2020; Manickam et al. 2021; Vandersluis et al. 2020), there remains a gap between the actual diagnostic rate and the suspected true prevalence of hereditary hearing loss (Smith et al. 2005). Furthermore, the improvement in the diagnostic yield for hearing loss as well as other pediatric rare diseases seems only modestly increased when comparing srGS to ES (Wojcik et al. 2024). Identifying the underlying cause of SNHL in pediatric patients is critical to providing tailored, high-quality care. Determining the genetic etiology of SNHL facilitates accurate genetic counseling and recurrence counseling, provides valuable information on prognosis that may impact treatment decisions, and allows for the identification and appropriate follow-up for children with syndromic SNHL. Ongoing clinical trials for targeted gene therapies for DFNB9 further emphasize the utility of precision medicine in the care of pediatric SNHL patients and the potential impact of an accurate genetic diagnosis (Lv et al. 2024).

Given the relatively high prevalence of pediatric SNHL in the population, an increase in the overall genetic diagnostic rate is poised to result in many more diagnoses per year, in absolute terms. The *STRC* gene alone has a carrier frequency of approximately 1.5% in the general population, and accounts for around 15% of all cases hereditary hearing loss (Han et al. 2021; Perry et al. 2023; Shearer et al. 2014; Shubina-Oleinik et al. 2021; Sloan-Heggen et al. 2016). However, due to the *STRC* pseudogene and complexity of this genomic region, many commercial labs will only perform CNV analysis without sequencing or omit *STRC* from gene panels altogether. We posit that *STRC* is, therefore, meaningfully under-diagnosed. We performed lrGS for two unrelated probands with congenital mild-moderate SNHL and a heterozygous loss of *STRC*, resulting in a high *a priori* suspicion for a missing second *STRC* variant. lrGS confirmed that suspicion, resolving loss-of-function SNVs *in trans* to a CNV for both probands. While less prevalent than *STRC-, OTOA-*mediated hearing loss is likely underdiagnosed as well due to its locus in a segmental duplication. Indeed, an estimated 7% of the human genome consists of segmental duplications that are highly mutable (Vollger et al. 2023; Vollger et al. 2022).

This work underscores the promise of lrGS to improve detection of variants in complex and difficult-to-resolve genomic regions and increase diagnostic yield. In this study, we selected a cohort of 19 probands with pediatric SNHL who had already had extensive genetic evaluation. We were able to identify four additional diagnoses and confirm a complex structural variant, leading to a diagnostic rate of 21.1% using lrGS. Emerging studies comparing the genetic diagnosis of rare disease using ES and srGS do not report a significant improvement in the yield of srGS over ES (Chung et al. 2023). SrGS remains more expensive than ES and more time-consuming to analyze. It is, in fact, not a part of routine clinical care to order srGS across most clinical indications, including pediatric SNHL. Sample sizes are small, but published rare disease studies using lrGS, including this one, hint at the potential for improved yield over other methodologies (Mahmoud et al. 2024; Miller et al. 2021; Pagnamenta et al. 2023; Xie et al. 2020).

LrGS, while not currently available on a clinical basis, offers a single test to identify SNV, CNVS, small insertion and deletions, tandem repeats, SVs, and methylation changes across both simple and complex genomic regions. In addition, this technology provides phasing information which may be advantageous for discrimination of pathogenic variation. For the patients in this small cohort, each had undergone extensive genetic testing prior to lrGS. While lrGS remains costly compared to other testing modalities, eliminating the use of other high-cost, low-yield tests such as srGS may ultimately be more beneficial to all stakeholders.

Limitations of our study include the small cohort size as well as the selection of probands for inclusion based on high *a priori* suspicion of a mendelian condition based on clinical characteristics. Future work should include a larger sample size of less highly selected patients with pediatric SNHL.

## Conclusions

Long read sequencing, while not in use clinically, has shown promise as a single test capable of detecting diverse genomic variant types in both simple and complex genomic regions. We describe four diagnoses identified in a cohort of 19 pediatric patients with SNHL who had undergone extensive nondiagnostic testing prior to lrGS, further highlighting the promise of this technology to significantly increase diagnostic rates.

## Data Availability

Most data needed to evaluate the conclusions in the paper are present in the paper and/or the Supplementary Materials. Additional data related to this paper may be requested from the authors.

## Statements and Declarations

## Acknowledgments

We thank the families for their participation.

## Funding

This work was funded by NIDCD K08 DC19716 and by Escuchar Sin Fronteras Foundation to AES with support from the Boston Children’s Hospital Rare Disease Cohort Initiative.

## Author contributions

AES and SER conceived the study. CTS and JMH contributed software. AES, CEF, CN, SER, WJR, and WS performed formal analyses that were validated by AES, CEF, SER, and WS. MZ, AP, CF, CL, EML, SER, and TS conducted the investigation process. AES, MK, and SER secured patient resources. AES, CEF, SER, and WS performed data curation. AES and SER wrote the original draft. AES, CEF, CN, EML, MK, SR, SER, WJR, and WS contributed to reviewing and editing. AES and SER prepared figures. AES, ME, MK, and SR provided supervision. SER, SR, and CEF contributed to project administration. All authors reviewed and approved of the manuscript.

## Compliance with ethical standards

### Conflict of interest

AES is the principal investigator of an investigator initiated clinical trial, Genetic Newborn Hearing Screening, funded by Akouos/Eli Lilly; the site principal investigator of the sponsored Phase I/II clinical trial *OTOF* gene therapy with Akous/Eli Lilly; and on the advisory board without compensation for Akous/Eli Lilly and Decibel/Akous. WJR, CN, JMH, CTS, CF, EML, CL, and MAE are employees and shareholders of PacBio. The other authors have no conflicts of interest to declare.

## Ethics approval

This study was approved by the institutional review board of Boston Children’s Hospital (IRB P00035179 and P00031494)

## Consent to participate

Written informed consent was obtained from all participants.

## Consent to publish

Consent to publish was obtained from all participants.

## Notes

### Funding Statement

This study was funded by NIDCD K08 DC19716 and by Escuchar Sin Fronteras Foundation to AES with support from the Boston Children's Hospital Rare Disease Cohort Initiative.

### Author Declarations

Ethics committee/IRB of Boston Children's Hospital gave ethical approval for this work. This work was approved under Boston Children's Hospital IRB protocols P00035179 and P00031494.

