## Supplementary material for "Long-Read Sequencing Increases Diagnostic Yield for Pediatric Sensorineural Hearing Loss": Supp. table 1 and supp. table 2

**Supplemental table 1**. Clinical and genetic data of long read genome sequencing cohort

| PacBio ID | Race | Ethnicity | Laterality | Sex | Onset | Symmetry | SNHL Severity (described) | SNHL Severity (summarized) | Family History | Other features | Single Gene (year performed) | Panel (year performed) | ES (year performed) | srGS (year performed) | lrGS | Other | Variants identified prior to lrGS (non-diagnostic) (variant- zygosity, ACMG Classification) | Long-Read Finding ( Variant - zygosity, ACMG classification) |
|---|---|---|---|---|---|---|---|---|---|---|---|---|---|---|---|---|---|---|
| BCH-01 | White | Not hispanic or latino | Bilateral | Female | Congenital | Asymmetric | Moderate rising to normal | Mild-Moderate | No | No | Yes | | Yes | Yes | Yes | | None | MITF; chr3:69977069_70380310inv (403.2 kb copy number neutral inversion) - het VUS |
| BCH-02 | White | Not hispanic or latino | Bilateral | Female | Congenital | Symmetric | Moderate to moderately-severe | Mild-Moderate | No | No | | Yes | Yes | Yes | Yes | | MYO7A:c.3476G>T (p.Gly1159Val) - het LP ; OTOF:c.1930G>A (p.Val644Ile) - het, VUS | OTOA; chr16:21747267_21748038 x1 (het del of 771 bp) - het P; NM_144672.3:c.2654A>G, p.His885Arg - het VUS |
| BCH-03 | White | Not hispanic or latino | Bilateral | Male | Congenital | Asymmetric | Slight right; moderate to prodound left | Severe-Profound | No | No | | | Yes | Yes | Yes | | HARS2:c.125T>G, p.L42* (NM_012208.2) - het, LP | None |
| BCH-04 | White | Not hispanic or latino | Bilateral | Female | Unknown | Symmetric | Mild to moderate | Mild-Moderate | No | No | | | Yes | Yes | Yes | | None | None |
| BCH-05 | White | Not hispanic or latino | Bilateral | Female | Postlingual | Asymmetric | Right moderate to severe; left slight to severe | Severe-Profound | No | No | | | Yes | Yes | Yes | | SLC26A4:c.1334T>G, p.Leu445Trp (NM_000441.2) - het, P | None |
| BCH-06 | White | Not hispanic or latino | Bilateral | Female | Congenital | Symmetric | Mild to moderate | Mild-Moderate | Yes | No | | | Yes | Yes | Yes | | STRC/CATSPER2 full gene deletion - het, P | STRC; NM_153700.2:c.1228C>T, p.Gln410* - het P; chr15:43891305_43941059x1 (STRC/CATSPER2 deletion) - het, P |
| BCH-07 | Asian | Not hispanic or latino | Bilateral | Male | Congenital | Symmetric | Moderate to severe cookie-bite, progressive | Severe-Profound | Yes | No | Yes | Yes | Yes | Yes | Yes | | GJB2:c.670A>C, p.Lys224Gln - het VUS; STRC:c.(?_3499-60)_(3557+31_?)del - het, P | None |
| BCH-08 | White | Not hispanic or latino | Bilateral | Male | Congenital | Symmetric | Moderate to moderately-severe | Severe-Profound | No | Hemolytic anemia | | | Yes | Yes | Yes | | None | None |
| BCH-09 | White | Not hispanic or latino | Bilateral | Female | Congenital | Symmetric | Profound | Severe-Profound | No | Retinal dystrophy | | Yes | Yes | Yes | Yes | CMA | None | None |
| BCH-10 | White | Not hispanic or latino | Bilateral | Male | Prelingual | Symmetric | Mild to moderately severe | Mild-Moderate | No | Global developmental delay | | | Yes | Yes | Yes | | None | None |
| BCH-11 | White | Not hispanic or latino | Bilateral | Female | Postlingual | Symmetric | Slight to moderate | Mild-Moderate | No | No | | | Yes | Yes | Yes | | None | None |
| BCH-12 | Unknown | Unknown | Bilateral | Female | Postlingual | Symmetric | Moderate to severe, progressive | Severe-Profound | No | No | | | Yes | | Yes | | None | None |
| BCH-13 | White | Not hispanic or latino | Bilateral | Female | Congenital | Symmetric | Severe rising to normal | Severe-Profound | Yes | No | | Yes | Yes | Yes | Yes | | GJB2:c.35del, p.Gly12fs* - het, P | None |
| BCH-14 | White | Hispanic or Latino | Unilateral | Female | Congenital | Unilateral | Profound | Severe-Profound | No | Bilateral enlarged vestibular aqeuduct | | | Yes | Yes | Yes | | None | None |
| BCH-15 | White | Not hispanic or latino | Bilateral | Female | Congenital | Symmetric | Mild to moderate | Mild-Moderate | No | vision, nystagmus | | | Yes | Yes | Yes | | USH2A:c.7475C>T, p.S2492L - het, LP | None |
| BCH-17 | White | Not hispanic or latino | Bilateral | Male | Unknown | Symmetric | Normal to severe | Severe-Profound | No | Hematuria | | | Yes | Yes | Yes | | USH2A:c.2276G>T, Cys759Phe (NM 206933.2) - het, P | None |
| BCH-18 | White | Not hispanic or latino | Bilateral | Female | Congenital | Symmetric | Severe to profound | Severe-Profound | No | No | Yes | | Yes | Yes | Yes | | None | None |
| BCH-19 | White | Not hispanic or latino | Bilateral | Male | Congenital | Symmetric | Mild to moderate | Mild-Moderate | No | No | | | Yes | Yes | Yes | | STRC/CATSPER2 full gene deletion - het, P | STRC; NM_153700.2:c.3217C>T - het, P; chr15:43890333_43940887x1 (STRC/CATSPER2 deletion) - het, P |
| BCH-20 | White | Not hispanic or latino | Bilateral | Female | Prelingual | Symmetric | Normal to mild | Mild-Moderate | Yes | No | | | Yes | Yes | Yes | | None | None |

**Supplemental table 2**. Coverage, read lenth, and number of varinats for short read exome sequencing (srES), short read genome sequencing (srGS), long read genome sequencing (lrGS).
Coverage = average read depth across loci; SNV = single nucleotide variant; SV = structural variant (>=50 bp)

| | srES | | | | srGS | | | | lrGS | | | |
|---|---|---|---|---|---|---|---|---|---|---|---|---|
| | **coverage** | **read length** | **SNV numb** | **SV number** | **coverage** | **read length** | **SNV numb** | **SV number** | **coverage** | **read length** | **SNV numb** | **SV number** |
| BCH-01 | 77.48 | 150 | 122829 | 72 | 51.08 | 150 | 5169563 | 12857 | 29.48 | 14975 | 5533276 | 23001 |
| BCH-02 | 118.97 | 150 | 124465 | 84 | 50.77 | 150 | 5035593 | 11261 | 30.84591 | 13314 | 5402271 | 21760 |
| BCH-03 | 90.63 | 150 | 125908 | 103 | 83.07 | 150 | 5163020 | 15639 | 29.46273 | 10901 | 5502079 | 23049 |
| BCH-04 | 86.93 | 150 | 119053 | 88 | 59.12 | 150 | 5074396 | 13851 | 23.51227 | 15048 | 5314161 | 19838 |
| BCH-05 | 124.05 | 150 | 121555 | 111 | 60.39 | 150 | 5054729 | 13753 | 29.14591 | 14752 | 5403655 | 21917 |
| BCH-06 | 87.94 | 150 | 119381 | 67 | 65.45 | 150 | 5028027 | 13910 | 28.63636 | 11700 | 5369081 | 22375 |
| BCH-07 | 54.44 | 150 | 119791 | 67 | 35.02 | 150 | 5027734 | 9768 | 31.31409 | 12176 | 5415720 | 22292 |
| BCH-08 | 55.88 | 150 | 111317 | 39 | | | | | 26.60273 | 13442 | 5372976 | 21486 |
| BCH-09 | 47.48 | 150 | 109565 | 83 | 28.71 | 150 | 4933372 | 9463 | 33.19045 | 15497 | 5322993 | 21922 |
| BCH-10 | 45.52 | 150 | 111855 | 66 | 42.26 | 150 | 5032567 | 11417 | 29.98409 | 13673 | 5386528 | 21565 |
| BCH-11 | 64.96 | 150 | 106362 | 49 | 50.53 | 150 | 5107243 | 12693 | 30.73273 | 14473 | 5467099 | 22619 |
| BCH-12 | 115.95 | 150 | 122261 | 115 | 51.89 | 150 | 5052255 | 12252 | 33.44545 | 14664 | 5430416 | 22097 |
| BCH-13 | 88.64 | 150 | 119855 | 83 | 43.41 | 150 | 5004984 | 12063 | 34 | 14009 | 5369919 | 22232 |
| BCH-14 | 65.71 | 150 | 108328 | 70 | 65.23 | 150 | 5022620 | 13345 | 30.46773 | 14692 | 5379248 | 22549 |
| BCH-15 | 60.9 | 150 | 115172 | 74 | 48.5 | 150 | 5006746 | 12599 | 24.19773 | 11913 | 5329772 | 22031 |
| BCH-17 | 60.31 | 150 | 111836 | 63 | 41.79 | 150 | 4951446 | 10025 | 28.78091 | 13003 | 5305533 | 21433 |
| BCH-18 | 55.22 | 150 | 115549 | 81 | 37.39 | 150 | 5034185 | 11120 | 28.92136 | 13315 | 5386525 | 21606 |
| BCH-19 | 46.25 | 150 | 122616 | 88 | 51.78 | 150 | 5013213 | 13187 | 20.08727 | 12400 | 5307152 | 21619 |
| BCH-20 | 87.69 | 150 | 121666 | 78 | 50.17 | 150 | 5137538 | 12287 | 30.87227 | 13539 | 5482230 | 22169 |
